# Advancing Cardiovascular Disease Diagnosis with an Interpretable and Responsible AI Framework

**DOI:** 10.1101/2025.06.17.25329798

**Authors:** Kazi Sakib Hasan, Irfan Sadi Dhrubo

**Affiliations:** School of Data and Sciences, BRAC University, Kha 224, Bir Uttam Rafiqul Islam Ave, Dhaka 1212, Bangladesh

## Abstract

Cardiovascular disease (CVD) remains a leading global health threat, responsible for one in five deaths worldwide. Early detection is critical to mitigate morbidity and mortality, yet traditional diagnostic methods often rely on reactive clinical assessments, missing opportunities for preventive intervention. In this study, a machine learning (ML) ecosystem is developed to enhance CVD diagnosis through two key approaches: (1) an early warning system using non-clinical, self-reported features for accessible risk stratification, and (2) specialized diagnostic models integrating clinical and non-clinical data. The framework leverages advanced ML techniques, including tabular neural networks (TabNet, TabPFN) and ensemble methods (XGBoost, Random Forest), validated on multi-regional datasets. Shapley Additive Explanations (SHAP) analysis identified ECG-related features as dominant predictors of CVD risk, with ST-segment slope (+0.93) and ST depression (+0.63) exhibiting the strongest effects. Counterfactual explanations from the non-clinical model further revealed actionable preventive measures: reducing exercise-induced angina and chest pain severity, alongside increasing exercise heart rate, could shift predictions from diseased to healthy, highlighting the model’s utility for lifestyle interventions. To address ethical and clinical trustworthiness, interpretability tools (SHAP, counterfactuals), fairness mitigation (FairLearn), and uncertainty quantification (Bayesian Neural Networks) are incorporated. Causal inference identified key predictors and their Average Treatment Effects (ATEs) such as exercise-induced angina (ATE: 0.36) and ST slope (ATE: 0.33), informing a hybrid ensemble model that achieved 89% accuracy while reducing dimensionality. The system aligns with FDA Good ML Practices and EU Trustworthy AI guidelines, offering a scalable solution for early detection and equitable diagnosis.

## 1 Introduction

Cardiovascular disease (CVD) is among the most serious health conditions, claiming one life every 33 seconds worldwide. It is also the leading cause of death for men, women, and individuals across most racial and ethnic groups [1]. In 2022, approximately 702,880 deaths were attributed to CVD, accounting for 1 in every 5 deaths [2]. Within the same year, nearly 1 in 5 deaths among adults under the age of 65 was caused by CVD [1]. It has been estimated that about 5% of adults above the age of 20 are affected by coronary artery disease. Reports from the CDC further indicate that approximately 1 in 5 heart attacks are silent, meaning patients are unaware of the damage sustained [3]. Because CVD can present with both symptomatic and asymptomatic characteristics, the disease often remains undiagnosed, which increases the risk of fatality. CVD typically affects the heart and blood vessels, manifesting as narrowed arteries, congenital structural problems, or malfunctioning heart valves. Mortality rates vary across sex, race, and ethnicity, highlighting the influence of lifestyle factors on disease prevalence [4]. Risk-enhancing lifestyle behaviors include smoking, poor diet, excessive alcohol consumption, and physical inactivity. In addition, medical conditions such as hypertension, diabetes, and hypercholesterolemia serve as major risk factors [3]. Environmental factors also play a role; for example, air pollution has been identified by the World Health Organization as a contributor to cardiovascular disease [5]. The complex interplay of these factors, combined with the frequent asymptomatic nature of CVD, often results in delayed or missed diagnoses. Such delays contribute to disease progression, increased healthcare costs, and, in severe cases, premature death. Heart conditions with lower prevalence may remain undiagnosed due to lack of familiarity or lower clinical suspicion, especially in pediatric practice. Misclassification of heart murmurs is also common in developing countries. Furthermore, organ dysfunction resulting from cardiovascular instability has been strongly associated with delayed or inaccurate diagnoses, leading to heightened morbidity and mortality [6]. For these reasons, early detection and preventive strategies are essential in reducing the burden of CVD.

Early identification of risk factors allows timely interventions through targeted medication and lifestyle modification, thereby preventing the progression of disease. It also reduces the need for expensive and invasive treatments in later stages. Moreover, early detection mitigates the risks associated with misdiagnosis, ensuring that conditions are identified before they advance silently. In essence, proactive diagnosis provides an opportunity to manage risk factors at the earliest phase, thereby limiting the severity of outcomes. Before examining early diagnostic strategies, it is important to consider traditional diagnostic approaches. Conventional cardiovascular workflows rely on established clinical protocols, where physicians gather information through physical examination, patient history, and diagnostic tests. These tests typically include electrocardiograms (ECGs), blood pressure measurements, cholesterol profiling, and stress tests. In advanced cases, invasive procedures such as angiography or fluoroscopy are employed. While these methods provide valuable insights, they are often resource-intensive, requiring costly equipment, specialized personnel, and significant time. Moreover, they are predominantly reactive, being deployed only after symptoms become apparent [7]. This limits opportunities for early detection, particularly in asymptomatic or atypical cases. Additionally, traditional methods rely on generalized risk models, which may not adequately account for individual variations in lifestyle, genetics, or environmental exposure, resulting in limited personalization in diagnosis and treatment planning. Machine learning (ML) offers a transformative alternative by enabling data-driven, personalized, and scalable diagnostic support. Trained on large datasets such as the UCI Heart Disease dataset, ML algorithms can uncover complex nonlinear relationships across multiple patient variables, patterns that might otherwise be overlooked in conventional assessments. These models not only improve diagnostic accuracy but also predict disease risk before the onset of symptoms. Importantly, retraining ML systems on non-invasive or self-reported data enhances accessibility and affordability. Furthermore, integration with wearables, lifestyle trackers, and electronic health records enables continuous monitoring and timely risk alerts. This shift supports low-cost, remote, and proactive care delivery, which is particularly valuable in underserved regions. While ML is not intended to replace clinical judgment, it functions as a powerful tool to augment decision-making, prioritize high-risk patients, reduce diagnostic delays, and enable more precise interventions.

Despite significant progress, current AI-based diagnostic systems for CVD continue to face several critical limitations. First, most existing models are diagnosis-oriented and seldom provide early warnings based on non-clinical or self-reported features, leaving a crucial gap in preventive care. Second, although AI adoption in healthcare is increasing, key issues such as interpretability, fairness, and predictive uncertainty are often neglected, reducing clinical trust and reliability. Third, the predominance of correlation-driven approaches has limited the incorporation of causal inference, thereby constraining model generalizability. Finally, advanced tabular neural networks (e.g., TabNet, TabPFN), which are particularly well-suited for healthcare data, remain largely unexplored in the context of CVD prediction.

To address these challenges, we propose a comprehensive, ethically aligned AI framework for CVD risk assessment with the following key contributions:

1. An early warning system that leverages non-clinical and self-reported features suitable for deployment via mobile health applications or community screening kiosks for accessible, cost-effective risk stratification.
2. Integration of interpretability tools (e.g., SHAP, counterfactual explanations), fairness mitigation (e.g., FairLearn), and uncertainty estimation (e.g., Bayesian neural networks, TabPFN).
3. Use of causal inference to identify mechanistic disease drivers beyond correlation.
4. Extensive benchmarking of advanced tabular neural networks against ensemble learning methods.

This framework is designed in alignment with regulatory guidelines from both the FDA and the European Union, with the goal of delivering a scalable, equitable, and clinically actionable solution for cardiovascular care.

## 2 Related Work

Machine learning (ML) and deep learning (DL) methods have been widely applied CVD prediction, with studies exploring feature selection, model optimization, and ensemble techniques. Despite strong performance across many works, challenges persist in generalizability, interpretability, and ethical deployment.

### 2.1 Traditional Machine Learning and Ensemble Models

Qadri et al. [8] proposed the Principal Component Heart Failure (PCHF) method, reducing dimensionality to eight features and reporting 100% accuracy using a Decision Tree—an outcome suggestive of overfitting. Kumar et al. [15] evaluated classical models on the UCI Heart Disease dataset and identified logistic regression as the most reliable. These studies underscore the utility of feature selection and simple classifiers but provide limited discussion on external validity and ethical considerations. Subramani et al. [11] introduced a stacking ensemble using IoT-based inputs, achieving 96% accuracy. Rohan et al. [14] compared 21 classifiers with 11 feature selection methods, identifying XGBoost as the top model (F1-score: 98%). Although ensemble methods consistently yield high accuracy, most works focus narrowly on performance metrics while overlooking transparency and fairness. Mohan et al. [22] applied Decision Tree entropy for feature extraction and achieved 0.887 accuracy using a Random Forest–Linear Regression hybrid. Ali et al. [23] reported perfect accuracy (1.0) with DT and RFC, raising concerns about potential overfitting. Bhatt et al. [24] conducted a more extensive evaluation involving DT, RF, XGBoost, and MLP with GridSearch tuning and Huang clustering, reaching an average cross-validation accuracy of 0.8707. Although diverse in methodology, these works tend to prioritize optimization over clinical interpretability.

### 2.2 Deep Learning Models and Feature Engineering

Deep neural networks (DNNs) have shown strong capacity for capturing nonlinear patterns in CVD data. Almazroi et al. [10] developed a Keras-based dense neural network that outperformed ensemble methods. Saeed et al. [12] combined DNNs with SelectKBest and SMOTE, reporting accuracies up to 99%. However, these studies provide limited attention to explainability. Al-Alshaikh et al. [9] proposed a hybrid system integrating genetic algorithms, recursive feature elimination, and a convolutional neural network optimized using adaptive elephant herd strategies. The approach delivered high recall (96.2%) but emphasized the need for broader real-world validation to improve generalization.

### 2.3 Innovative Neural and Bio-Inspired Techniques

Novel architectures and bio-inspired optimizers have been explored for CVD prediction. Nandy et al. [16] introduced the Swarm-Artificial Neural Network (Swarm-ANN), achieving 95.78% accuracy by using heuristic-driven weight adjustments. While effective, interpretability remained limited. Eleyan et al. [13] developed Rhythmi, a CNN-based mobile diagnostic tool for ECG analysis, reporting over 98% accuracy. Although promising for real-time diagnosis, the work relied on a relatively small dataset and focused primarily on signal data rather than tabular clinical datasets commonly found in EHRs.

### 2.4 Recent Advances in Limited Data Integration

Recent studies have examined ML and DL approaches for integrating heterogeneous or limited clinical datasets. Mehdi et al. [29] distinguished normal and impaired cardiomyocytes using sarcomere transients and calcium kinetics, achieving AUC values of 0.94–0.95 through an ensemble classifier. Building on multimodal integration, Mehdi et al. [30] proposed a multifidelity ML framework for myocardial infarction diagnosis that combined simulation-based low-fidelity data with limited human CMR data, improving Dice scores from 0.39 to 0.72. Their later work [31] estimated myocardial mechanical properties directly from anatomical and hemodynamic features, reporting high predictive accuracy (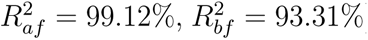). Pressure–volume loading emerged as the most informative feature set. Collectively, these studies highlight the promise of multimodal integration and highlight gaps in interpretability, fairness, and uncertainty quantification—areas critical for developing clinically trustworthy AI systems.

## 3 Methodology

Multiple models were developed for clinical and non-clinical applications. Non-clinical modeling used three ensemble methods, while clinical modeling employed tabular neural networks, a causality-guided voting classifier (CGEVC), Bayesian neural networks, and the ensemble baselines. The framework incorporated interpretability and uncertainty quantification.

A diverse set of machine learning and deep learning methods was chosen to align with regulatory priorities (FDA, EU AI Act) emphasizing interpretability, uncertainty, causality, and robustness. Ensembles (Random Forest, XGBoost, LightGBM) served as strong baselines, tabular neural networks (TabNet, TabPFN, FT-Transformer) offered interpretable generalization for small datasets, Bayesian networks quantified uncertainty, and CGEVC prioritized causally relevant features. This multimodel framework enabled systematic evaluation of robustness, interpretability, fairness, and uncertainty, while identifying models suitable for real-world deployment. Reproducibility guidelines are provided in the GitHub repository [27]. The overall workflow is illustrated in Figure 1.

**Figure 1:**
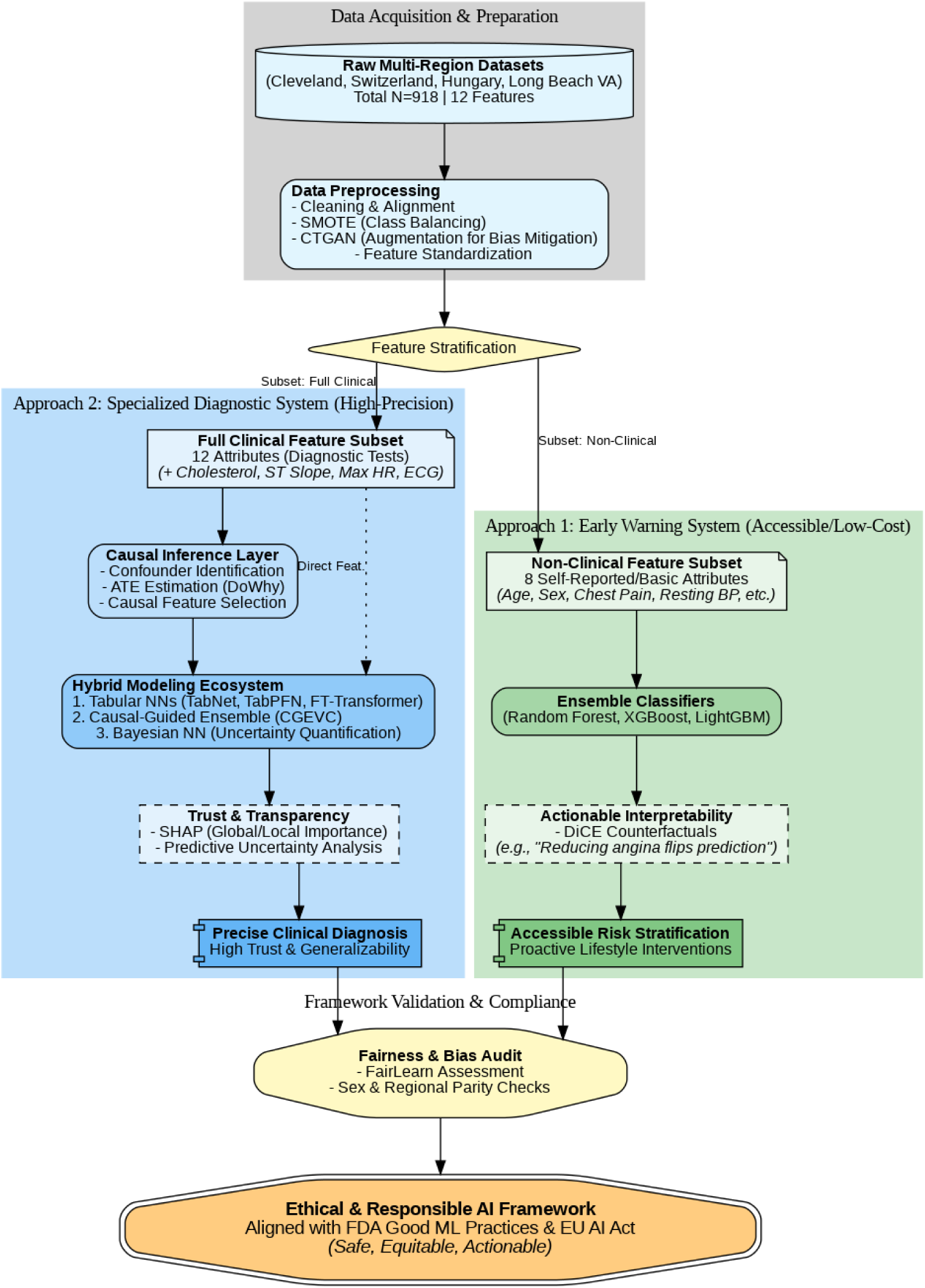
This diagram illustrates the methodology for the two key approaches described in the study. (1) Non-Clinical Stream: Designed for low-resource settings, this stream uses accessible patient history features to train interpretability-focused models (Random Forest, XGBoost) that suggest lifestyle changes via DiCE counterfactuals. (2) Clinical Stream: Designed for medical settings, this stream incorporates diagnostic test results (e.g., ST slope, Serum Cholesterol) into causality-guided and probabilistic neural architectures (TabPFN, BNN) to ensure clinical reliability. The shared evaluation layer ensures both streams meet ethical standards for fairness and transparency.

### 3.1 Dataset Information

Two datasets from Kaggle were utilized, both derived from the UCI Heart Disease dataset [28]. To avoid ambiguity, they are distinguished as follows:

- **Cleveland:** A Kaggle-provided dataset containing only the Cleveland Clinic data, with 303 instances and 14 features. This dataset has been widely adopted in heart disease prediction research.
- **Multi-Region-Combined:** A merged dataset comprising records from five sources: Cleveland, Switzerland, Long Beach VA, Hungary, and Statslog (Kaggle-provided). The initial version contained 1,190 instances.

The combined dataset exhibited several inconsistencies that required preprocessing:

1. The features ca (number of major vessels colored by fluoroscopy) and thal (thalassemia) were missing from the Switzerland, Long Beach VA, and Hungary subsets. To maintain feature consistency, these attributes were removed from the Cleveland and Statslog subsets as well.
2. Column names in the Statslog dataset were inconsistent with the Cleveland schema. These were standardized to ensure alignment.
3. Detailed inspection revealed that all 272 rows in Statslog were duplicates of rows from the Cleveland dataset, differing only in order. These duplicates were removed.

After preprocessing, the final **Multi-Region**dataset contained 918 unique instances aggregated from four sources: Cleveland, Switzerland, Long Beach VA, and Hungary. Table 1 summarizes the datasets, their sizes, and their differences, providing an overview of the data sources used in this study. Figure 2 shows how each dataset is derived and used in the research.

**Figure 2:**
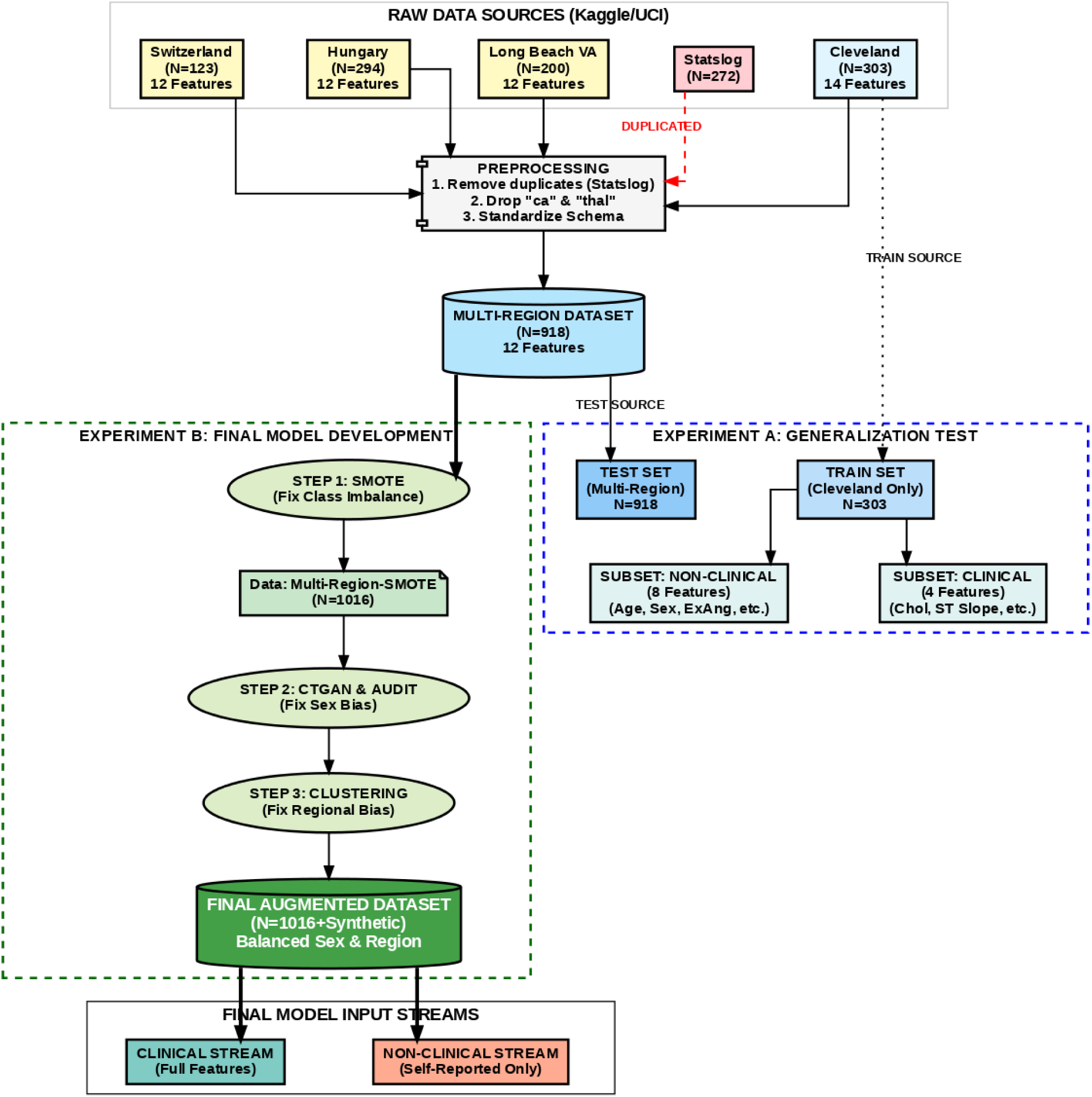
The diagram illustrates the consolidation of five raw data sources (Cleveland, Hungary, Switzerland, Long Beach VA, and Statslog) into a unified Multi-Region dataset (*N* = 918). Preprocessing involved removing the Statslog subset due to duplication and dropping inconsistent features (ca, thal) to ensure schema alignment. The workflow bifurcates into two experimental phases: **(A) Generalization Testing**, where models trained solely on Cleveland (*N* = 303) are evaluated on the combined dataset; and **(B) Final Model Development**, which employs a multi-stage bias mitigation pipeline. This pipeline integrates **SMOTE** for class balancing (*N* = 1016), **CTGAN** for demographic parity (Sex), and unsupervised **Clustering** to address regional bias. Finally, features are stratified into **Clinical** and **Non-Clinical** streams to support the framework’s dual-model architecture.

**Table 1:** Summary of the datasets used in this study. The table categorizes data into raw sources, the unified multi-region set, and the specific subsets derived for generalization testing and final model development.

| Dataset | Instances | Features | Description / Notes |
| --- | --- | --- | --- |
| ***Raw Data Sources*** |  |  |  |
| Cleveland | 303 | 14 | Original Cleveland data (Kaggle). |
| Statslog | 272 | 14 | Removed (Duplicates of Cleveland). |
| Switzerland | 123 | 12 | Missing `ca` and `thal` features. |
| Long Beach VA | 200 | 12 | Missing `ca` and `thal` features. |
| Hungary | 294 | 12 | Missing `ca` and `thal` features. |
| ***Merged Data*** |  |  |  |
| **Multi-Region** | **918** | **12** | Aggregation of Cleveland + 3 regional datasets. |
| ***Experiment A: Generalization Test Subsets*** |  |  |  |
| Cleveland-Clinical | 303 | 4 | Train set for clinical modeling. |
| Cleveland-Non-clinical | 303 | 8 | Train set for non-clinical modeling. |
| Multi-Region-Clinical | 918 | 4 | Test set for clinical generalization. |
| Multi-Region-Non-clinical | 918 | 8 | Test set for non-clinical generalization. |
| ***Experiment B: Final Modeling (Augmented)*** |  |  |  |
| Multi-Region-SMOTE | 1016 | 12 | Balanced via SMOTE. Basis for final models. |
| Clinical Dataset | 1016 | 4 | Final input for TabNet, TabPFN, etc. |
| Non-clinical Dataset | 1016 | 8 | Final input for Random Forest, Ensembles. |

#### 3.1.1 Generalization Test

To evaluate generalization, models were first trained on the Cleveland dataset and then tested on the Multi-Region dataset. For both datasets, features were divided into two categories:

- **Clinical features:** serum cholesterol, resting electrocardiographic result, ST depression induced by exercise relative to rest, and slope of the peak exercise ST segment.
- **Non-clinical features:** all remaining attributes.

This division yielded four additional subsets: Cleveland-Clinical, Cleveland-Non-Clinical, Multi-Region-Clinical, and Multi-Region-Non-Clinical. Models trained on non-clinical subsets provide early-warning predictions, whereas clinical subsets incorporate diagnostic test outcomes.

Three ensemble models (XGBoost, LightGBM, and Random Forest) were trained on the Cleveland-Clinical and Cleveland-Non-Clinical datasets, and subsequently evaluated on the corresponding Multi-Region subsets to assess generalization ability. Performance with accuracy above 0.80 and ROC-AUC above 0.85 was regarded as evidence of strong generalization.

### 3.2 Modeling on Multi-Regional Data

After confirming generalization from Cleveland to Multi-Region data, final model development was conducted using the Multi-Region dataset. To mitigate slight class imbalance, the Synthetic Minority Oversampling Technique (SMOTE) was applied.

Figure 3 illustrates the class distribution before and after applying SMOTE. SMOTE generates synthetic samples for the minority class to balance imbalanced datasets. Given that generalization has already been established, the use of SMOTE does not raise ethical concerns regarding data leakage. Unlike simple duplication, SMOTE creates new instances by interpolating between existing minority class samples, ensuring that the augmented data introduces meaningful variability while preserving the original data distribution.

**Figure 3:**
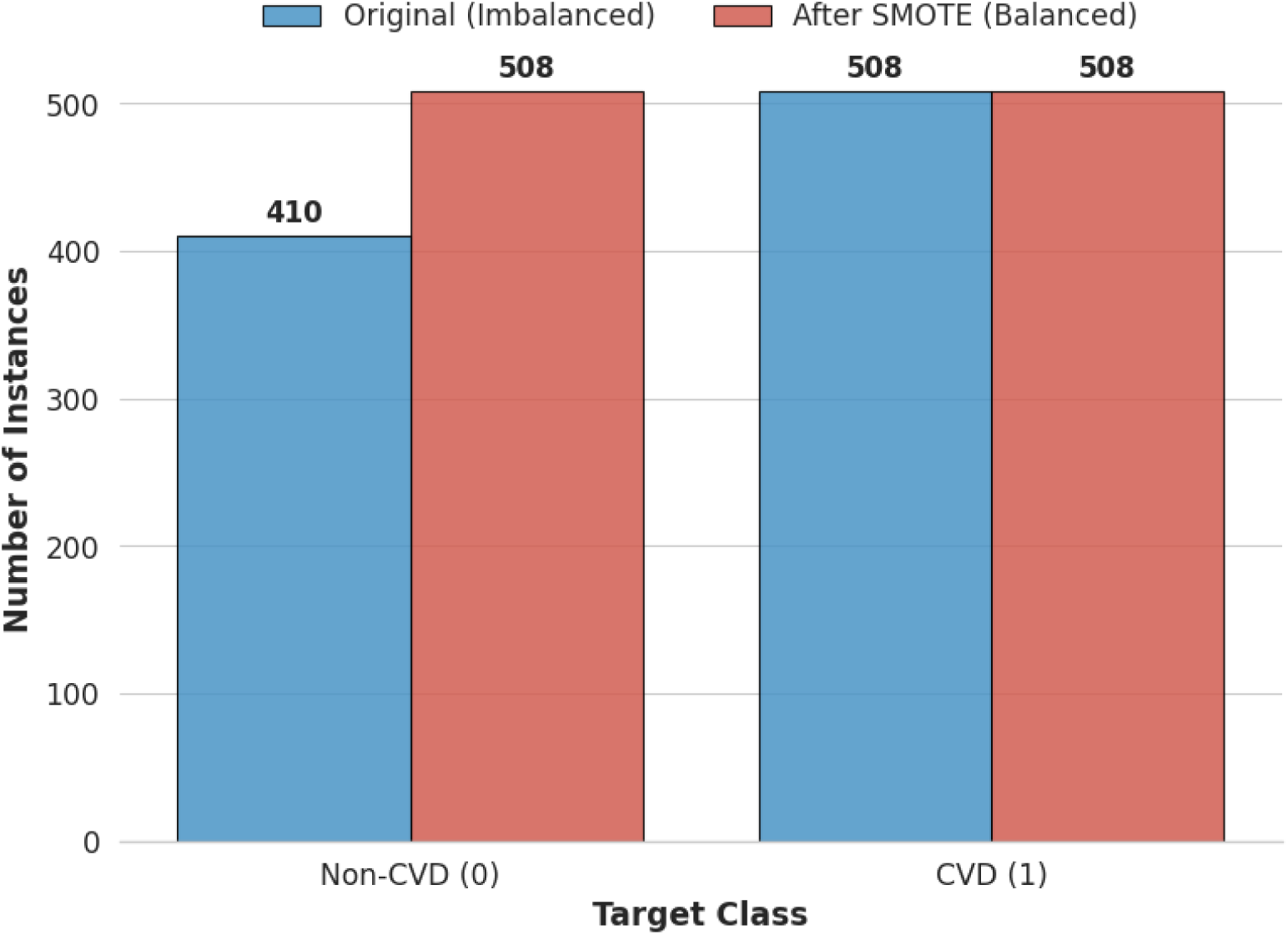
Class distribution of the target variable (heart disease presence) in the multi-regional dataset before and after applying the SMOTE, demonstrating the effective balancing of the dataset.

Given a minority class sample **x***_i_*, a synthetic sample **x̃** is generated as:

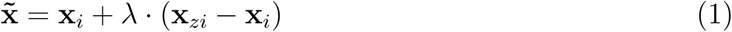

where **x***_i_* is a minority class instance, **x***_zi_* is one of the *k* nearest neighbors of **x***_i_*, and *λ* ∼ U(0, 1) is a random number drawn from a uniform distribution. This process generates synthetic data points along line segments connecting minority class neighbors, reducing overfitting and improving model generalization on imbalanced datasets. The oversampling procedure was applied once on the combined dataset, after which clinical features were removed to create separate clinical and non-clinical subsets. The three ensemble models were then trained and evaluated on both subsets.

While the initial performance was promising, the bias report from FairLearn indicated a tendency for the models to predict CVD more frequently for men, while predicting most women as healthy. This pattern may reflect the natural epidemiology of CVD, as men generally exhibit higher incidence rates. However, the observed bias still raises ethical concerns. To address this, an additional experiment was conducted using data augmentation with a Conditional Tabular Generative Adversarial Network (CTGAN). Synthetic instances were generated to equalize the frequencies of diseased men and women, as well as healthy men and women. This experiment demonstrates that balancing the dataset can mitigate bias without requiring complex methods such as adversarial debiasing or algorithmic regularization.

Importantly, CTGAN was not applied to improve model performance in this study. The generative process does not introduce data leakage, as CTGAN is specifically designed for tabular data and effectively handles imbalanced, mixed-type datasets (both continuous and categorical features). By using a conditional generator, the model captures the distribution of minority classes and rare categories more accurately.

Integrating a conditional generator into a GAN architecture involves addressing three key challenges. First, an appropriate representation of the condition must be devised and provided as input. Second, the generated rows must faithfully preserve the specified condition. Third, the conditional generator must learn the true conditional distribution of the real data, ensuring realistic and representative synthetic samples.

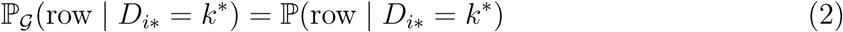

so that we can reconstruct the original distribution as

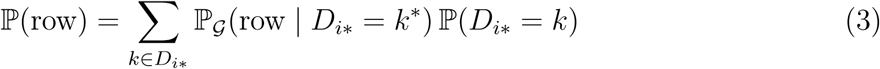

To effectively model continuous columns, CTGAN employs mode-specific normalization using Variational Gaussian Mixture Models (VGM). For discrete columns, conditioning vectors **c** are sampled to guide generation, ensuring better representation of underrepresented categories [25]. This conditional mechanism enables CTGAN to generate diverse and realistic synthetic tabular data, particularly in class-imbalanced scenarios. After applying CTGAN, the three ensemble models were retrained on the augmented dataset, resulting in balanced bias metrics across genders.

To fully address demographic bias, it was also necessary to evaluate potential regional bias. Although the final model does not include region as a feature, since real-world users are unlikely to correspond only to Switzerland, Long Beach VA, Cleveland, or Hungary, the goal was to generalize the model across all regions rather than rely solely on external validation. To achieve this, theoretical regions were extracted from the dataset using unsupervised learning. K-Means clustering, combined with Principal Component Analysis (PCA), was applied to identify four regions. The unsupervised approach produced a silhouette score of 0.21, and the resulting regional frequencies closely mirrored those of the original dataset, as illustrated in Figure 4.

**Figure 4:**
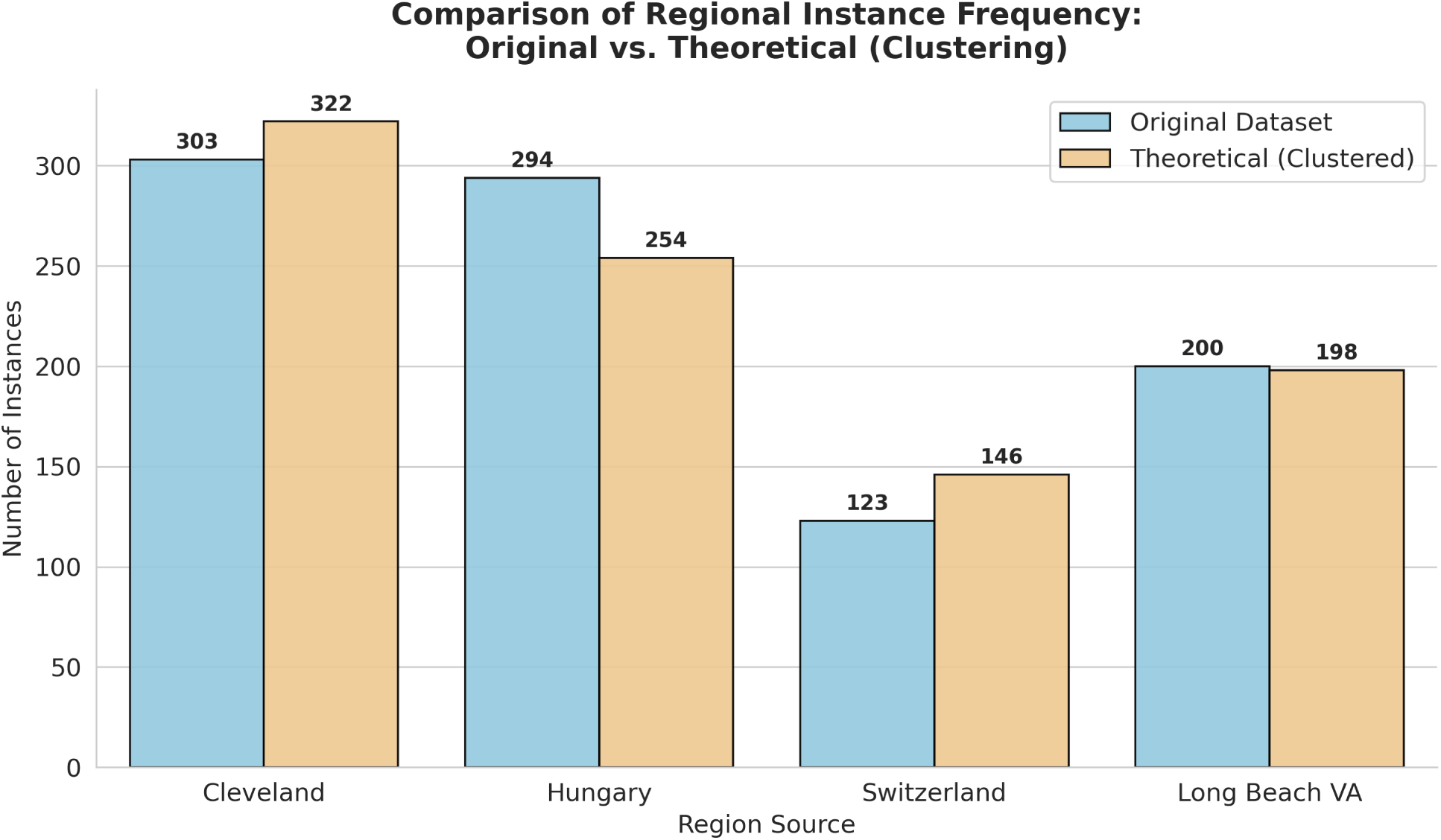
Distributional validation of the unsupervised regional clustering. The blue bars represent the actual instance counts from the four source datasets (Cleveland, Hungary, Switzerland, Long Beach VA), while the orange bars represent the clusters identified via PCA and K-Means. The close alignment in frequency counts (Silhouette Score: 0.21) confirms that the unsupervised theoretical regions effectively reconstruct the original geographical sources, validating their use for assessing regional bias.

The three ensemble models were subsequently trained on the dataset augmented with the additional theoretical region feature, and bias metrics were evaluated. The analysis revealed notable insights, which motivated the use of agglomerative clustering instead of K-Means with PCA, as detailed in the Results section. Following this, the combined dataset with SMOTE and CTGAN (excluding the region feature) was used to develop three tabular neural network models: TabNet, FT-Transformer, and TabPFN. A Bayesian Neural Network (BNN) employing Monte Carlo Dropout was also implemented for uncertainty quantification. Additionally, a causality-guided voting classifier, composed of XGBoost, LightGBM, Random Forest, K-Nearest Neighbors, and Logistic Regression, was developed to enhance prediction accuracy. Prior to training the neural network models, the dataset was standardized using scikit-learn’s StandardScaler(), which performs z-score normalization on all features. The voting classifier was trained using only features with an Average Treatment Effect (ATE) of at least 0.1 on CVD diagnosis.

### 3.3 Model Selection

We employed three ensemble models, three tabular neural networks, and a Bayesian Neural Network (BNN). Non-clinical models use only ensembles; clinical models use all architectures.

#### 3.3.1 Ensemble Models

##### XGBoost

Boosted trees optimized via a regularized objective; well-suited for mixed tabular data and tuned using Bayesian Optimization.

##### Random Forest

Averaged predictions from many bootstrapped trees; reduces variance and overfitting in small clinical datasets.

##### LightGBM

Optimized tree learner using GOSS and EFB for efficient training; hyperparameters tuned via Optuna.

#### 3.3.2 Tabular Neural Networks

##### TabNet

Sequential attention selects features sparsely, providing interpretability and strong performance on mixed data.

##### TabPFN

A pretrained Bayesian few-shot learner that approximates posterior predictions without tuning, ideal for small datasets.

##### FT-Transformer

Applies self-attention over feature tokens; default hyperparameters used to prevent overfitting on small samples.

#### 3.3.3 Bayesian Neural Network (BNN)

##### BNN with MC Dropout

Learns weight distributions and estimates predictive uncertainty via multiple stochastic forward passes, beneficial for high-risk clinical prediction.

#### 3.3.4 Causal-Guided Ensemble Voting Classifier (CGEVC)

A causal-guided ensemble classifier is designed to enhance the robustness and interpretability of cardiovascular disease (CVD) prediction. The workflow integrates confounder discovery, causal effect estimation, and ensemble learning. The pipeline is described below:

##### 1. Identifying Potential Confounders

To estimate the causal effect of each feature (treatment variable) on the target (CVD diagnosis), the potential confounders are firstly identified using a data-driven approach. For each feature *T_i_*:

1. Remove the target variable *Y* from the dataset.
2. Use a Random Forest Classifier (RFC) to predict *T_i_* using the remaining features **X** \ *T_i_*.
3. Extract the feature importances from RFC and select the subset **Z***_i_* ⊂ **X** \ *T_i_* such that the cumulative importance satisfies:

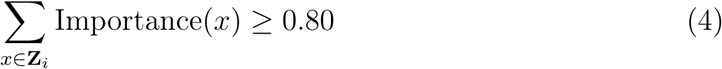

The selected features **Z***_i_* are considered potential confounders for estimating the causal effect of *T_i_*on *Y* .

##### 2. Estimating Average Treatment Effect (ATE)

The backdoor criterion is used from causal inference to estimate the Average Treatment Effect (ATE) of each treatment *T_i_* on the outcome *Y* , conditioned on the identified confounders **Z***_i_*. According to the backdoor adjustment formula:

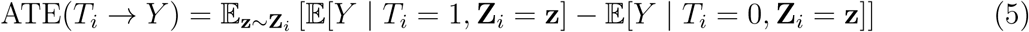

This is implemented via the DoWhy framework with the backdoor.linear_regression estimator, which assumes the linear model:

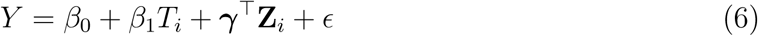

The coefficient *β*_1_ provides the estimated causal effect of *T_i_* on *Y* , while controlling for confounders **Z***_i_*.

##### 3. Causal-Guided Feature Selection and Ensemble Classification

Features with statistically significant (0.1) ATE values are selected to train a soft voting ensemble classifier composed of:

- Random Forest Classifier (RFC)
- XGBoost
- LightGBM
- *k*-Nearest Neighbors (KNN)
- Logistic Regression (LR)

The final prediction *ŷ* is the weighted average of predicted class probabilities:

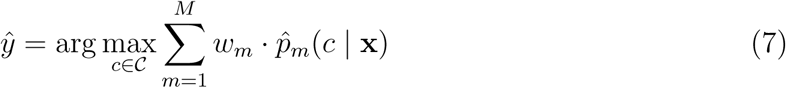

where *w_m_*is the weight of model *m*, and *p̂_m_* is the predicted probability for class *c*.

This pipeline integrates causal inference into model training, helping to focus on variables that have meaningful causal influence on the target. This improves interpretability, reduces bias from spurious correlations, and enhances performance.

## 4 Results and Discussion

Table 2 details the classification performance of the ensemble models across both experimental setups. It highlights that the application of SMOTE and multi-regional training data significantly enhanced model precision and recall compared to the baseline generalization tests.

**Table 2:**
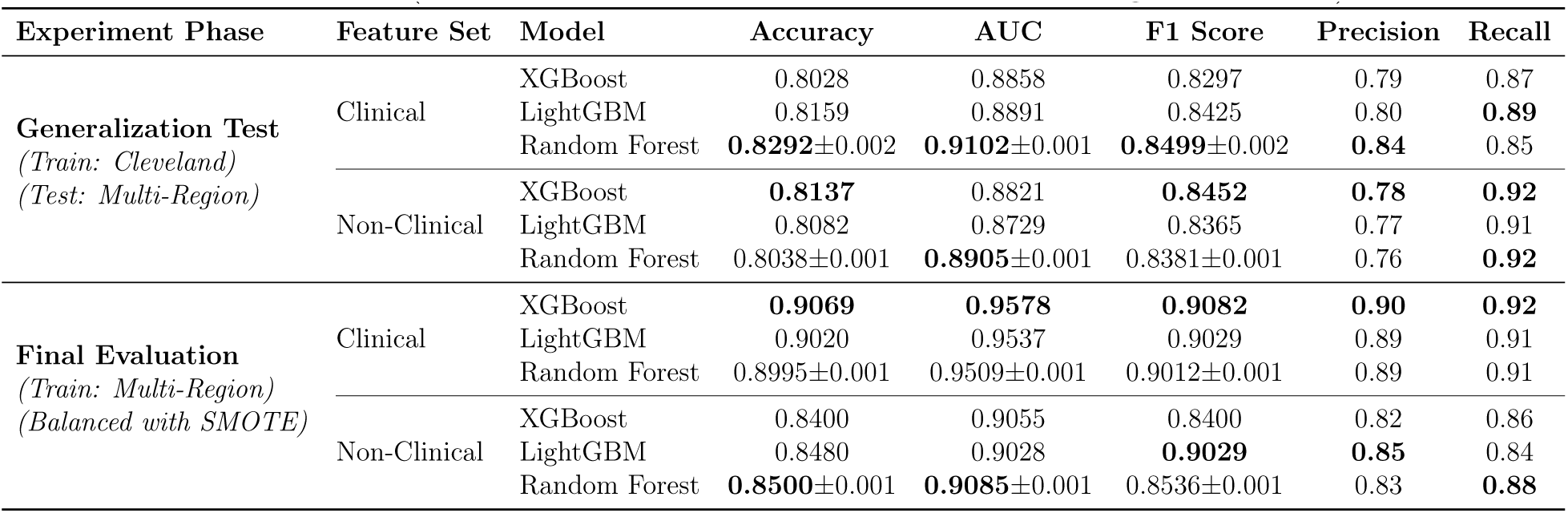
Performance comparison across training phases. The table aggregates results for the **Generalization Test** (models trained on Cleveland, tested on Multi-Region) and the **Final Evaluation** (models trained on the balanced Multi-Region dataset).

Figure 5 visualizes that most blue bars (healthy) are clustered at low probabilities (0–0.3) for the imbalanced dataset. Many red bars (disease) are pushed toward lower probabilities (0.2–0.4), instead of being clearly separated. This suggests the model (XGBoost trained on clinical dataset) struggles to predict disease correctly, indicating it’s biased toward the majority class (healthy) due to class imbalance. Conversely, the red bars (disease) are spread across higher probabilities (0.5–1.0), while blue bars (healthy) remain mostly at low probabilities (0–0.3) for the balanced dataset. There is better separation between healthy and disease, meaning the model can distinguish the classes more confidently. So, SMOTE helped the model learn patterns of the minority class, improving prediction for disease cases.

**Figure 5:**
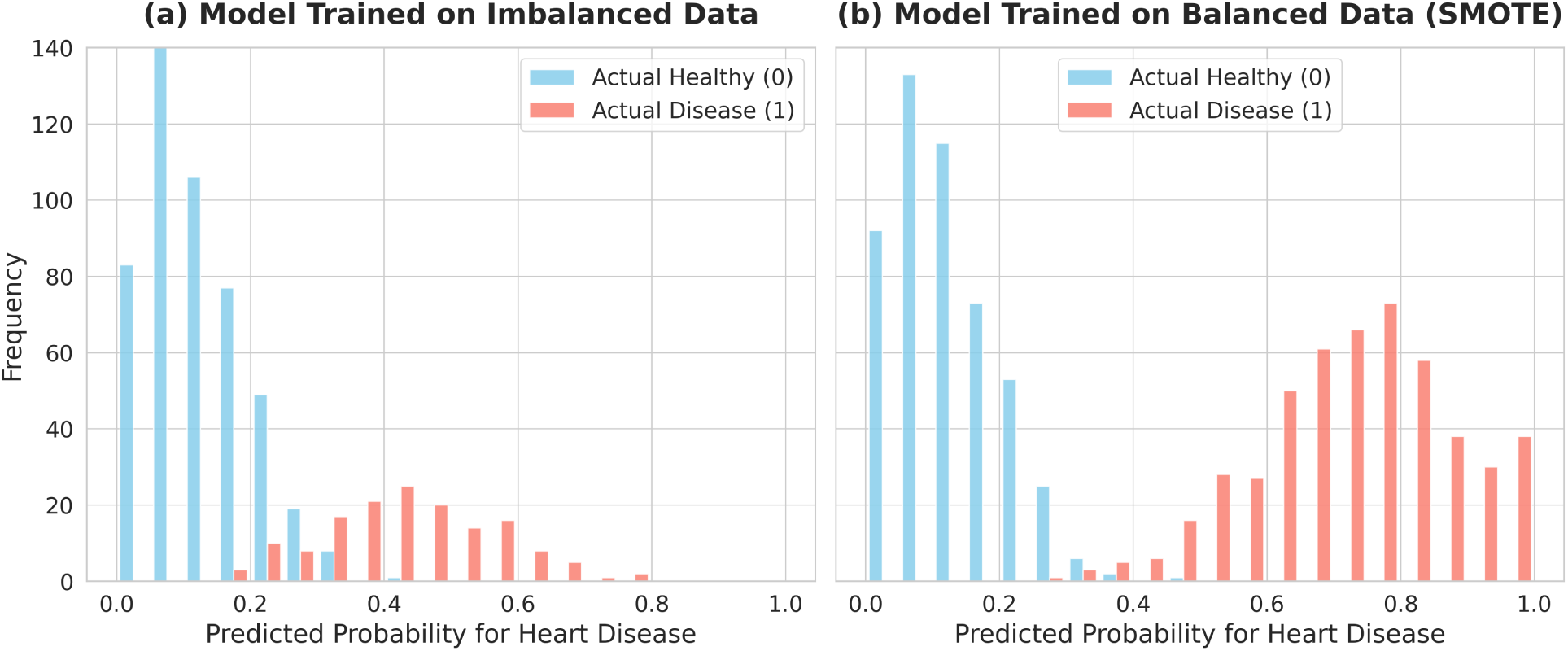
Histograms of predicted probabilities for the XGBoost clinical model. (a) visualizes the distribution on the original imbalanced dataset, and the SMOTE-balanced dataset is visualized in (b). The balanced dataset shows improved separation between healthy (blue) and diseased (red) classes.

### 4.1 Fairness Metrics

From Table 3, Both clinical (XGBoost) and non-clinical (RF) models show noticeable disparities in sex-wise predictions. The selection rate for males is substantially higher (62% vs 22% for XGBoost, and 63.3% vs 22.7% for RF), which directly contributes to a demographic parity difference (DPD) of 0.40 in both cases. This implies that males are much more likely to be predicted as positive compared to females. Likewise, the demographic parity ratio (DPR) is only 0.35, far below the commonly accepted fairness threshold of 0.8 (the “80% rule” [26]), suggesting potential adverse impact.

**Table 3:**
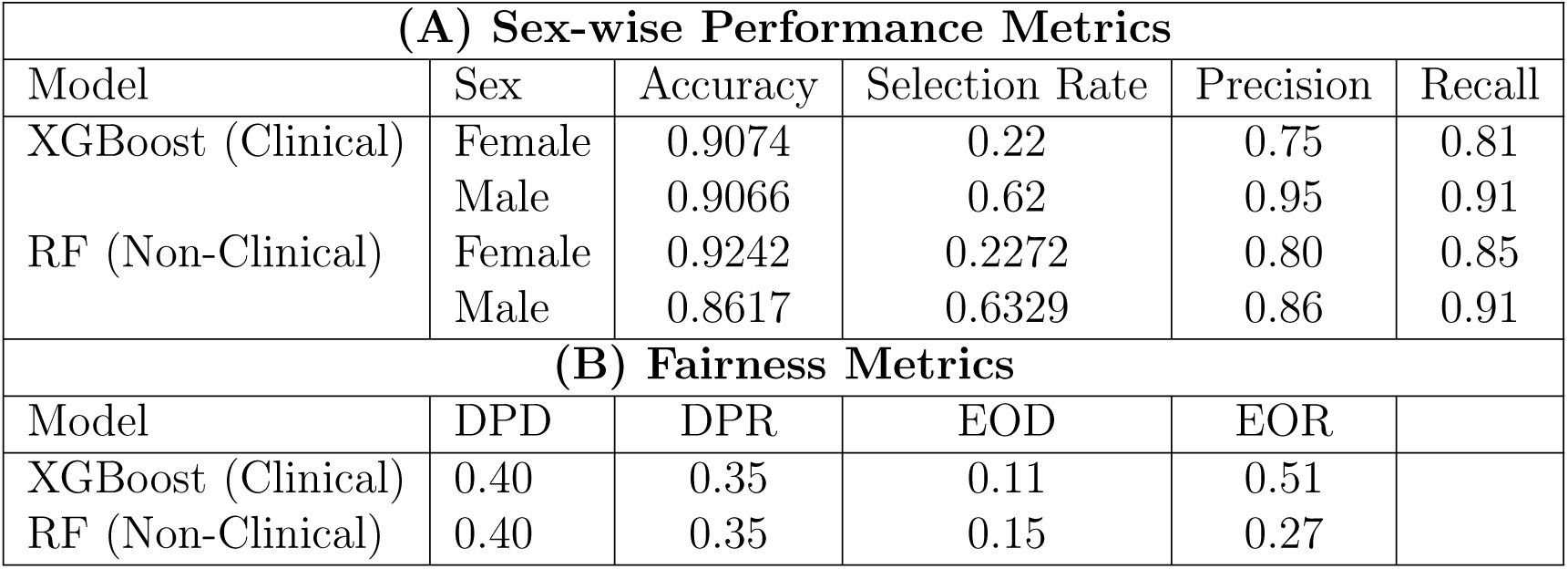
(A) Sex-wise performance metrics (Accuracy, Selection Rate, Precision, Recall) and (B) corresponding fairness metrics (Demographic Parity Difference/DPD, Demographic Parity Ratio/DPR, Equalized Odds Difference/EOD, Equalized Odds Ratio/EOR) for the primary clinical (XGBoost) and non-clinical (Random Forest) models, highlighting initial demographic disparities.

| (A) Sex-wise Performance Metrics |  |  |  |  |  |
| --- | --- | --- | --- | --- | --- |
| Model | Sex | Accuracy | Selection Rate | Precision | Recall |
| XGBoost (Clinical) | Female | 0.9074 | 0.22 | 0.75 | 0.81 |
|  | Male | 0.9066 | 0.62 | 0.95 | 0.91 |
| RF (Non-Clinical) | Female | 0.9242 | 0.2272 | 0.80 | 0.85 |
|  | Male | 0.8617 | 0.6329 | 0.86 | 0.91 |
| (B) Fairness Metrics |  |  |  |  |  |
| Model | DPD | DPR | EOD | EOR |  |
| XGBoost (Clinical) | 0.40 | 0.35 | 0.11 | 0.51 |  |
| RF (Non-Clinical) | 0.40 | 0.35 | 0.15 | 0.27 |  |

Precision and recall values also reveal disparities: males tend to receive higher precision (0.95 for XGBoost, 0.86 for RF) while females exhibit lower precision but relatively balanced recall. Equalized odds differences (EOD) of 0.11 (XGBoost) and 0.15 (RF) further confirm discrepancies in true/false positive rates, with equalized odds ratios (EOR) well below 1 (0.51 and 0.27, respectively). Ideally, both should be close to 0 (difference) and 1 (ratio), respectively. However, these disparities may not solely reflect algorithmic bias but could also be linked to the underlying prevalence of heart disease by sex, as illustrated in Figure 6.

**Figure 6:**
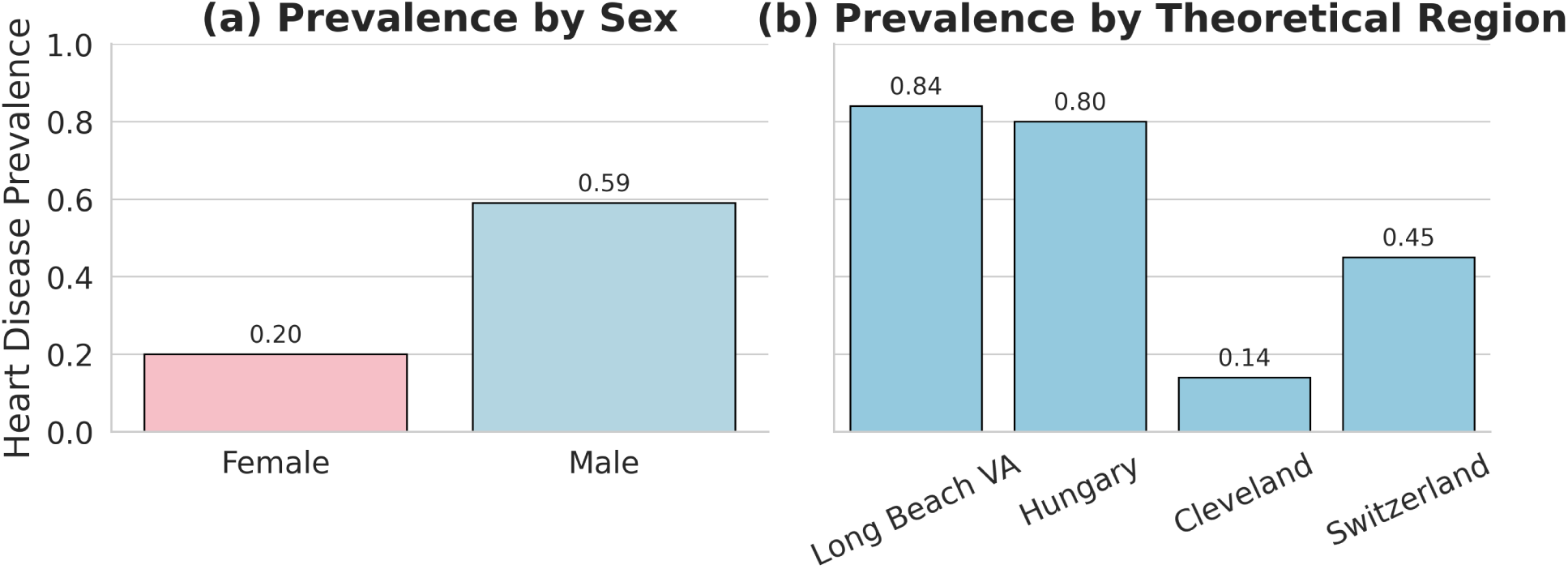
Bar chart illustrating the underlying prevalence of heart disease by sex and by the theoretically derived regions within the dataset, providing context for observed biases in model predictions.

Figure 6 also illustrates that, beyond sex-based fairness, regional bias could also exist because of Cleveland’s low prevalence rate. The selection rate and recall of Cleveland (theoretical) were significantly lower compared to other clusters, for both clinical (0.13) and non-clinical (0.082) models. Such imbalance could be due to differences in heart disease prevalence across regions, as shown in Figure 6(b), or the limitations of the PCA + K-Means clustering approach.

To address this, agglomerative clustering was attempted, yielding four clusters with a silhouette score of 0.44. One cluster, however, contained only 8 instances and shared distributional similarity with another cluster. It indicates that one region (e.g., Cleveland) closely resembles to some other region (e.g., Longbeach VA), and therefore we can drop it to analyze disparities with more transparency. Hence, we retained three clusters instead of four and retrained the models, which improved balance in both performance and fairness metrics across regions. The final selection rates for the three regions were 0.52, 0.92, and 0.42 respectively, supporting the robustness and generalizability of the models under external validation.

#### 4.1.1 Bias Mitigation Results

After applying CTGAN and retraining the models, fairness metrics improved significantly.

For the clinical model XGBoost:

- Demographic parity difference = 0.05
- Demographic parity ratio = 0.91
- Equalized odds difference = 0.09
- Equalized odds ratio = 0.55

For the non-clinical model Random Forest Classifier:

- Demographic parity difference = 0.07
- Demographic parity ratio = 0.892
- Equalized odds difference = 0.089
- Equalized odds ratio = 0.715

The fairness evaluation of both models shows acceptable performance in terms of demographic parity than before. However, it reveals some concerns regarding equalized odds. The general model exhibits a demographic parity difference of 0.05 and a ratio of 0.91, indicating a minor disparity in selection rates between groups. Similarly, the Random Forest (non-clinical) model has a slightly higher demographic parity difference of 0.07 and a ratio of 0.892, both within commonly accepted fairness thresholds. However, equalized odds metrics are a bit problematic: the general model shows a difference of 0.09 and a low ratio of 0.55, suggesting an imbalance in error rates across demographic groups. The Random Forest model performs slightly better with an equalized odds difference of 0.089 and a ratio of 0.715, though still below the 0.8 fairness threshold. These results imply that while group-wise selection rates are relatively fair, there is disparity in how accurately the models treat different groups, warranting mitigation efforts focused on reducing outcome disparities. Future research may look into this further.

### 4.2 Model Interpretability and Counterfactuals

SHAP interpretations and counterfactual explanations were incorporated into both clinical and non-clinical models, enabling comprehensive global and local interpretability for each prediction. In clinical models, SHAP identified variables such as chest pain type, exercise-induced angina, and ST slope as critical contributors. In non-clinical models, behavioral and symptom-related features, including chest pain and heart rate response, were emphasized. Counterfactual explanations, generated using DiCE, complemented SHAP by illustrating minimal actionable changes capable of altering a prediction from diseased to non-diseased. These interpretability tools enhance transparency and model trustworthiness while providing clinicians and users with actionable insights into potential lifestyle or symptom-based interventions. This aligns with ethical AI principles and increases the suitability of the models for real-world healthcare deployment.

The SHAP summary plot and instance-level interpretation for the clinical model, shown in Figure 7(b), depict the global feature importance and their impact on XGBoost predictions. Features such as slope, oldpeak, and chest pain type exert the greatest influence, with wider distributions indicating stronger effects. Positive SHAP values (right side) increase predicted CVD risk, whereas negative values (left side) reduce risk estimates. The plot also indicates that age and sex have relatively smaller, yet consistent, impacts compared to other clinical features such as maximum heart rate and serum cholesterol.

**Figure 7:**
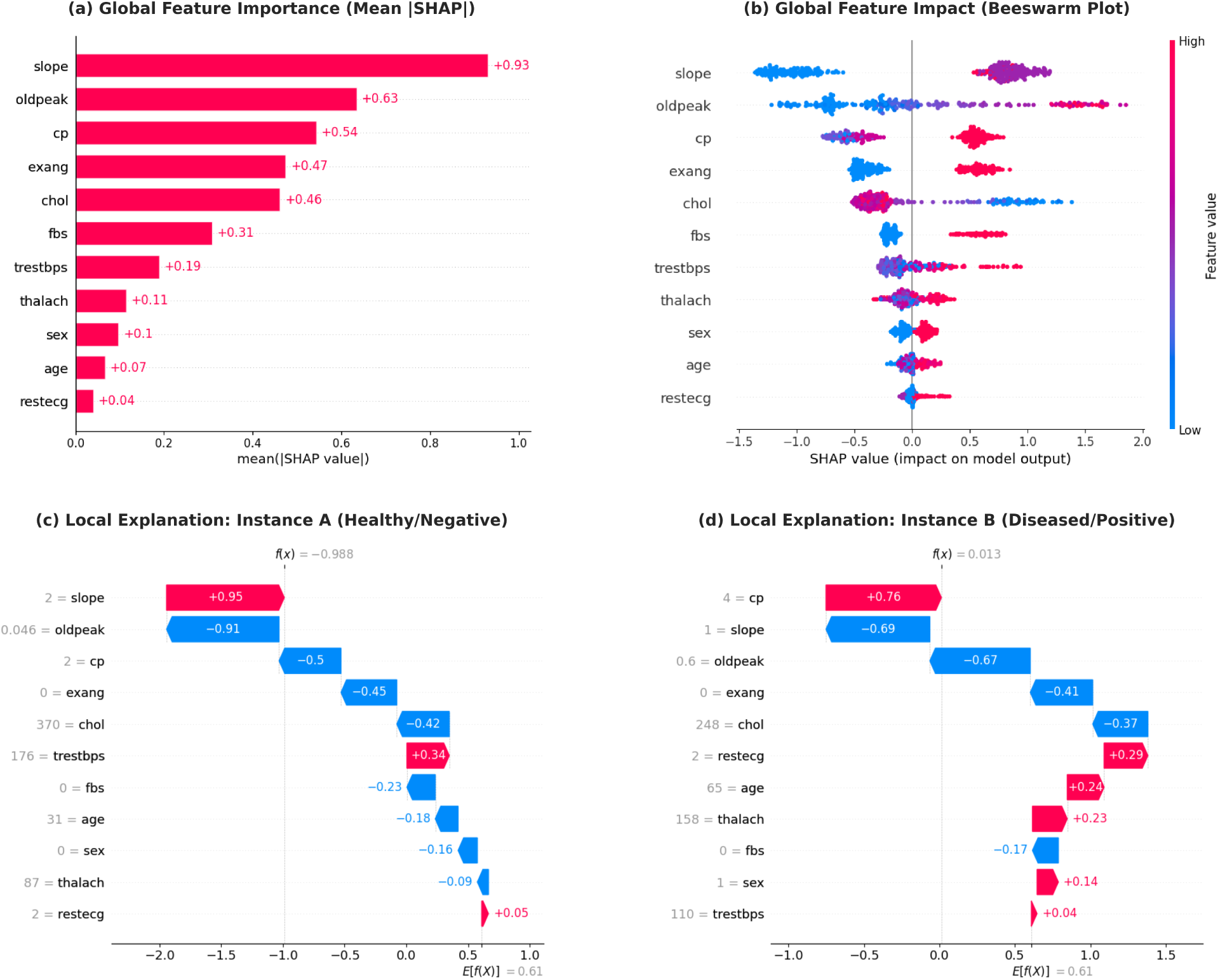
SHAP analysis for the XGBoost clinical model. (a) shows global feature importance, (b) illustrates the global feature impact (beeswarm plot) on model outcome, and the waterfall plot in (c) and (d) provides a local explanation for a specific instance predicted as healthy and diseased, respectively. These visualizations highlight the contribution of each feature to the prediction.

Figure 7(d) visualizes that the model’s base value, or average prediction across the dataset, was 0.61, and the prediction for this specific instance rose to approximately 0.623, crossing the classification threshold. The strongest contributor to the positive prediction was the chest pain type (cp = 4), typically associated with asymptomatic or severe angina, contributing +0.76 to the final score. Additional risk factors included abnormal resting ECG results (restecg = 2), older age (65), male sex, and a high maximum heart rate (thalach = 158). Despite several features that reduced the risk, such as a flat ST slope (slope = 1), low ST depression (oldpeak = 0.6), absence of exercise-induced angina (exang = 0), and near-normal cholesterol levels- the cumulative weight of high-risk indicators led the model to classify the patient as having heart disease. This example highlights the model’s ability to weigh clinical features in a nuanced manner, with significant emphasis on symptom severity and ECG abnormalities, aligning well with medical understanding. Similarly, Figure 7(c) shows similar interpretation for a healthy individual. Figure 7(a) reveals the global feature importance for the XGBoost CVD prediction model:

- **Top Influencers:**

- slope (ST-segment slope, +0.93) and oldpeak (ST depression, +0.63) are the strongest predictors, indicating ECG-related features dominate CVD risk.
- cp (chest pain type, +0.54) and exang (exercise-induced angina, +0.47) follow closely, highlighting the importance of clinical symptoms.
- **Moderate Contributors:**

- chol (cholesterol, +0.46) and fbs (fasting blood sugar, +0.31) show measurable but smaller impacts.
- **Minimal Influence:**

- Demographic factors (age +0.07, sex +0.10) and restecg (resting ECG, +0.04) have relatively low importance.

The model prioritizes direct cardiac indicators over traditional risk factors, with ECG-related features (slope, oldpeak) showing nearly twice the impact of cholesterol levels. This aligns with clinical intuition that direct physiological markers are more predictive for CVD diagnosis than demographic variables.

The local interpretation and mean SHAP values of **non-clinical model (RF)** is shown in Figure 8.

**Figure 8:**
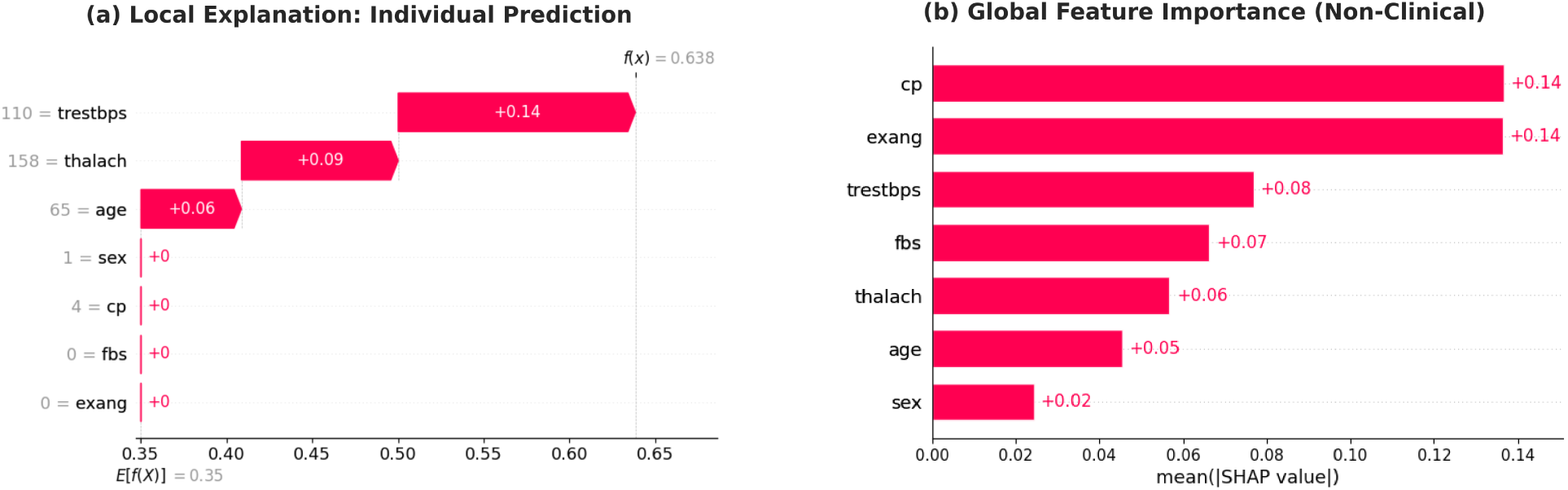
Interpretability analysis for the Non-Clinical Random Forest model. (a) Local waterfall plot explaining an individual prediction; basic health metrics like age and max heart rate (thalach) push the prediction toward disease, while the absence of exercise-induced angina (exang) lowers the risk. (b) Global feature importance (Mean |SHAP|) identifying chest pain type (cp) and exercise-induced angina (exang) as the most critical self-reported predictors for the early warning system.

The interpretation method for these plots are similar as XGBoost. Figure 8 shows that the individual’s age, max heart rate, and resting blood pressure are the major factors behind getting diagnosed as CVD.

Figure 9(a) presents the global feature importance derived from TabNet, highlighting the overall contribution of each variable to the model’s predictions. Figure 9(d) corresponds to a correctly predicted healthy individual, while Figure 9(c) represents a diseased case. These interpretations are made possible through TabNet’s intrinsic sparse attention mechanism, which enables the model to selectively focus on the most relevant features for each individual prediction. TabNet also supports SHAP interpretation, as shown in Figure 9(b).

**Figure 9:**
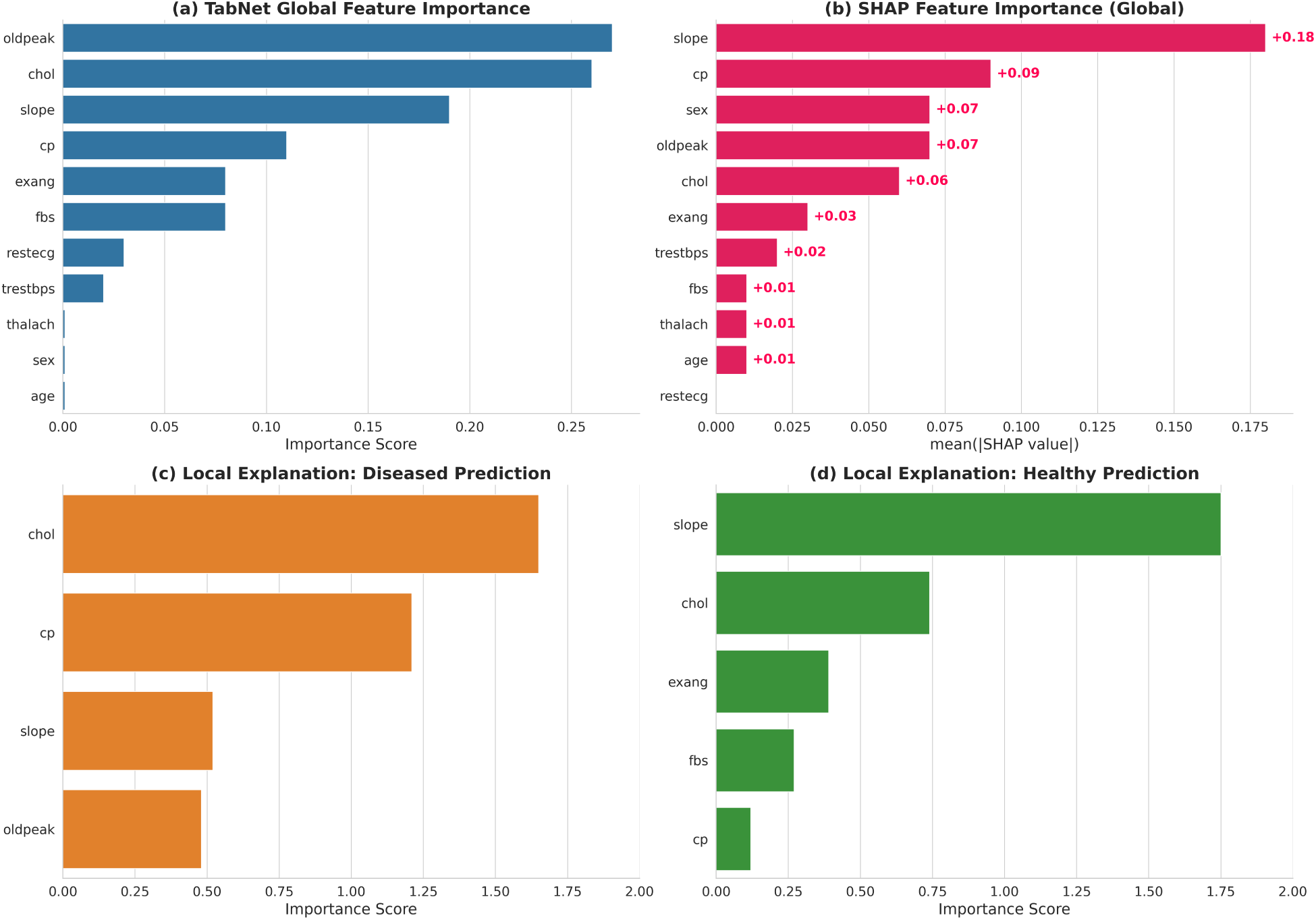
Interpretability of the TabNet model. (a) shows global feature importance from TabNet’s intrinsic sparse attention mechanism. (c) and (d) provide local explanations for a diseased and a healthy prediction, respectively. (b) shows a SHAP summary plot for TabNet.

### 4.3 Counterfactual Analysis

Counterfactual analysis was used to explore how slight changes in input features could alter the model’s prediction from **diseased** to **healthy**. By identifying minimal, plausible modifications such as reducing chest pain type or increasing ST slope, actionable insights are revealed that may inform personalized interventions. This approach enhances interpretability by showing not only why a prediction was made, but also how it could change.

Table 4 shows that, for the clinical model, two distinct pathways to a healthy prediction were identified. The values specified in parentheses correspond to the predicted probability of disease; a probability below 0.5 shifts the classification from ‘Disease’ to ‘Healthy’. The first pathway suggests a reduction in symptom severity—specifically, shifting chest pain presentation from asymptomatic to typical angina (cp: 4 → 1) and eliminating exercise-induced angina (exang: 1 → 0). The second pathway highlights the model’s reliance on physiological markers; normalizing the ST slope (slope: 3 → 1) and ST depression (oldpeak: 1.5 → 0.0) alone was sufficient to flip the prediction, even if other risk factors remained constant.

**Table 4:** Counterfactual explanations generated using DiCE. The table compares the original high-risk patient record (Age 38, Male, Resting BP 110) against generated counterfactuals. **Bold values** indicate the specific changes required to flip the prediction from Disease to Healthy. The values in parentheses in the Prediction column represent the model’s predicted probability of cardiovascular disease. Note that Age and Sex were held constant to ensure physiological plausibility. (*N/A indicates feature not used in that model* ).

| Model | Scenario | Key Features |  |  |  |  | Prediction |
| --- | --- | --- | --- | --- | --- | --- | --- |
|  |  | CP Type | Ex. Angina | ST Slope | ST Depr. | Max HR |  |
| **Clinical** | Original Patient | 4 | Yes (1) | 3 | 1.5 | 105 | Disease (0.91) |
|  | Counterfactual 1 | **1** | **No (0)** | 3 | 1.5 | 105 | **Healthy (0.38)** |
|  | Counterfactual 2 | 4 | Yes (1) | **1** | **0.0** | 105 | **Healthy (0.42)** |
| **Non-Clinical** | Original Patient | 4 | Yes (1) | N/A | N/A | 105 | Disease (0.86) |
|  | Counterfactual 1 | **2** | **No (0)** | N/A | N/A | 105 | **Healthy (0.45)** |
|  | Counterfactual 2 | 4 | **No (0)** | N/A | N/A | **183** | **Healthy (0.41)** |

Abbreviations: CP = Chest Pain, ST Depr. = ST Depression (oldpeak), Max HR = Maximum Heart Rate (thalach).

For the non-clinical Model, which relies on self-reported and basic health data, the counterfactuals provide actionable lifestyle targets. As shown in the bottom section of Table 4, increasing cardiovascular fitness—simulated by raising the maximum heart rate (thalach) from 105 to 183, or strictly eliminating exercise-induced angina resulted in a healthy classification.

### 4.4 Neural Networks Performance, Interpretation, and Uncertainty

This section focuses on the performance of neural network architectures.

Table 5 shows that among all the evaluated neural models, TabPFN outperforms others with the highest accuracy (90%), macro F1-score (0.90), and weighted F1-score (0.90). This consistency across all key metrics indicates its superior balance in handling both positive and negative classes effectively. The model’s performance reflects not only strong predictive ability but also generalizability across patient groups. CGEVC’s balanced performance similar to other models indicate that the causal factors were truly important features for CVD diagnosis. Also, due to filtering out features that have an ATE of less than 0.1, CGEVC successfully reduced the dimensionality while preserving diagnosis performance.

**Table 5:** Performance comparison of advanced neural models and the Causal-Guided Ensemble Voting Classifier (CGEVC) on the cardiovascular disease prediction task. TabPFN achieved the highest accuracy and F1-scores, while CGEVC maintained strong performance with reduced feature dimensionality.

| Model | Accuracy | Macro F1-Score | Weighted F1-Score |
| --- | --- | --- | --- |
| TabNet | 0.89 | 0.88 | 0.89 |
| FT-Transformer | 0.89 | 0.88 | 0.89 |
| TabPFN | **0.90** | **0.90** | **0.90** |
| BNN | 0.87 | 0.86 | 0.87 |
| CGEVC | 0.89 | 0.88 | 0.89 |

#### 4.4.1 Uncertainty Quantification

The outer histogram on Figure 10(a) illustrates the uncertainty distribution (measured as the standard deviation of predictive probabilities) for both CVD and non-CVD predictions using the Bayesian Neural Network.

**Figure 10:**
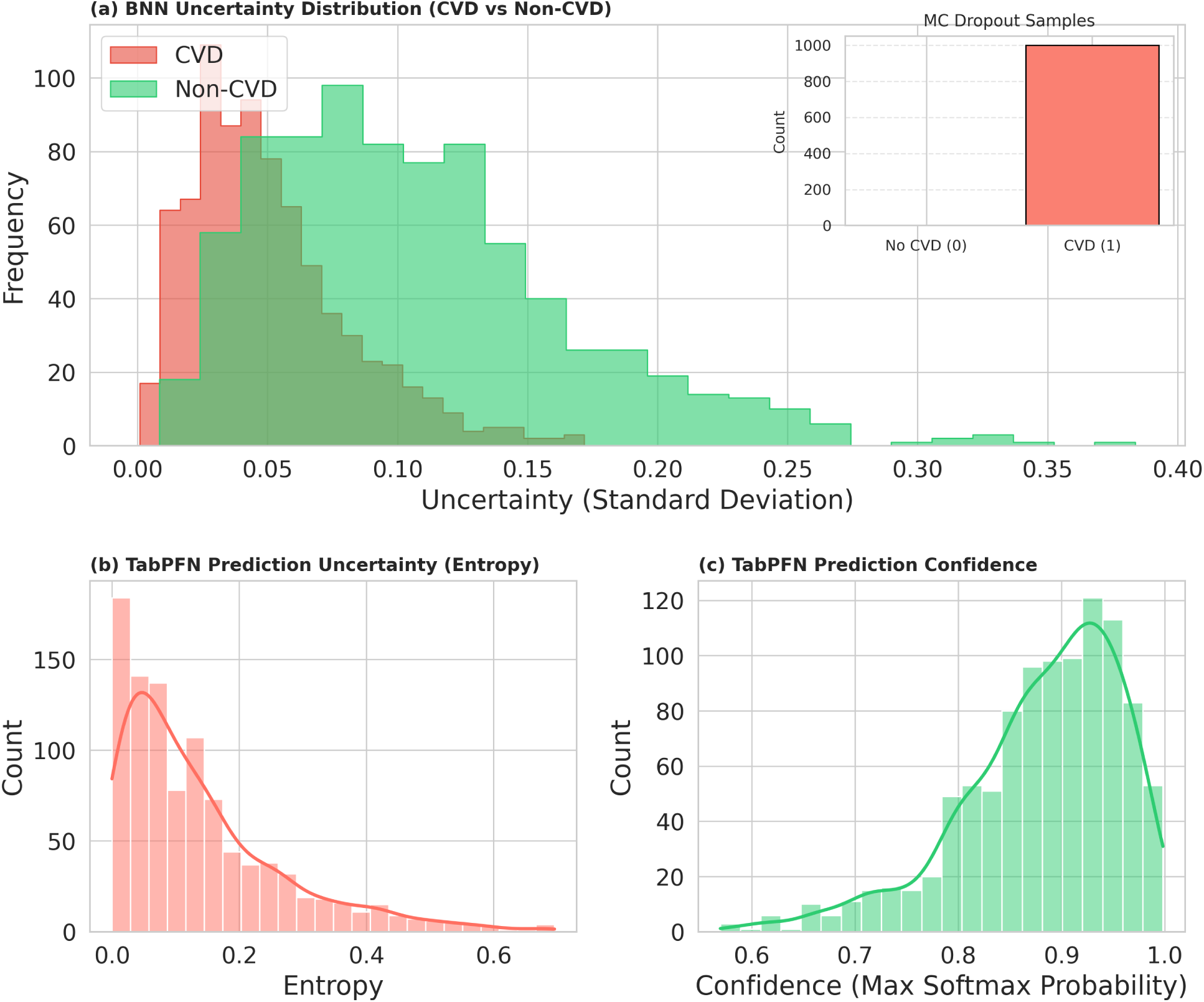
Uncertainty quantification for the Bayesian Neural Network and TabPFN. (a) shows the BNN’s uncertainty distribution (outer histogram) and prediction characteristics for a random instance (inner bar chart). (b) presents TabPFN’s predictive uncertainty via entropy and (c) shows confidence distributions.

Figure 10(a) visualizes in its outer histogram that, the majority of predictions, for both classes, exhibit low uncertainty, concentrated between 0 and 0.1, indicating the model was generally confident in its predictions. However, CVD predictions (in red) are slightly more concentrated in the very low uncertainty range (around 0.03–0.08), whereas non-CVD predictions (in green) display a slightly wider spread, with some extending up to 0.35. This may suggest the model is more certain when predicting diseased individuals than when ruling out disease. Nonetheless, both distributions taper off beyond 0.15, showing that high-uncertainty predictions are infrequent, which is a desirable trait for clinical reliability.

The inner bar chart of Figure 10(a) illustrates the prediction characteristics of the Bayesian Neural Network (BNN). Owing to its probabilistic nature, the BNN samples multiple weight configurations during inference, resulting in a distribution of predictions for each instance. A sample is classified as class 1 (diseased) if the mean of these predictions exceeds 0.5. A prediction count concentrated near the total number of Monte Carlo (MC) samples indicates high confidence, while broader distributions reflect greater uncertainty. This approach offers a principled and robust quantification of uncertainty, a distinctive capability of Bayesian models.

The uncertainty analysis for TabPFN is presented through two key metrics: entropy and prediction confidence. In the entropy histogram shown in Figure 10(b), we observe that most predictions are concentrated at very low entropy values (close to 0), indicating that the model often outputs highly confident and sharp class probabilities. However, there’s a notable tail extending up to 0.7, suggesting that a minority of predictions remain highly uncertain, potentially due to ambiguous or borderline cases. The confidence histogram in Figure 10(c) complements this by showing that the majority of predictions have a softmax maximum probability above 0.90, further confirming the model’s decisiveness on most samples. The sharp peak near 1.0 in the confidence plot supports the low-entropy pattern observed earlier. Together, these visualizations imply that TabPFN tends to be highly confident in its predictions, but retains the ability to flag uncertainty where necessary, which is essential for trustworthy AI in clinical settings.

### 4.5 Causal Inference

A clean summary and interpretation of causal inference results is provided in this section.

#### 4.5.1 Estimated Causal Effects (Average Treatment Effect - ATE)

In the context of causal inference frameworks (such as potential outcomes), the term ‘treatment’ refers to the intervention variable or exposure whose effect is being estimated, rather than a clinical medical therapy. In this analysis, each feature—including immutable characteristics such as Age and Sex, is mathematically modeled as a ‘treatment’ variable. This allows us to estimate the ATE of that specific feature on the probability of CVD diagnosis, while controlling for relevant confounders.

##### 1. Age

- ATE: 0.0045
- Confounders: oldpeak, thalach, trestbps, chol

Interpretation: Age has a small but positive causal effect on heart disease. As age increases, the likelihood of disease slightly rises. However, the effect is relatively modest, likely due to its indirect interaction through other clinical variables. Statistically, on average, one unit change in age results in 0.45% point increase in the probability of heart disease, assuming other confounders are held constant.

##### 2. Sex

- ATE: 0.1274
- Confounders: oldpeak, thalach, trestbps, age

Interpretation: Being male is associated with a higher risk of heart disease, which aligns with epidemiological findings. The effect is moderate, and appears independent of core vitals and exercise-induced markers. ATE indicates that one unit change in Sex (practically, male to female, or female to male) results in 12.74% point increase in the probability of heart disease, assuming the confounders are held constant.

##### 3. Chest Pain Type (cp)

- 0.1651
- Confounders: oldpeak, thalach, trestbps, chol, age

Interpretation: Chest pain type exerts a strong causal influence. This confirms its clinical importance as a diagnostic feature, especially when chest pain is atypical or exertional. Statistically, on average, a one-unit change in the cp (chest pain type) results in a 16.51% point increase in the probability of heart disease, assuming other confounders are held constant.

##### 4. Resting Blood Pressure (trestbps)

- ATE: 0.0011
- Confounders: oldpeak, thalach, chol, age

Interpretation: Despite its clinical relevance, resting blood pressure demonstrates a minimal causal effect on heart disease in this analysis. The ATE value suggests that, on average, a one-unit increase in resting blood pressure leads to a 0.11% point increase in the probability of heart disease, assuming confounders are held constant. Its effect may be mediated or overshadowed by stronger cardiovascular markers.

##### 5. Serum Cholesterol (chol)

- ATE: -0.00089
- Confounders: oldpeak, thalach, trestbps, age

Interpretation: Surprisingly, cholesterol exhibits a very weak negative causal effect on heart disease in this dataset. This could indicate that its influence is either non-linear, confounded, or not directly contributing in the presence of other variables. A one-unit increase in cholesterol is associated with a 0.089% point decrease in heart disease probability, assuming confounders remain fixed.

##### 6. Fasting Blood Sugar (fbs)

- ATE: 0.2566
- Confounders: thalach, trestbps, chol, age

Interpretation: Fasting blood sugar shows a strong positive causal relationship with heart disease, highlighting the impact of glucose metabolism and diabetes on cardiovascular risk. On average, a one-unit increase in fbs (indicating elevated blood sugar) increases the probability of heart disease by 25.66% points, holding other factors constant.

##### 7. Resting ECG (restecg)

- ATE: 0.0375
- Confounders: thalach, trestbps, chol, age

Interpretation: Resting ECG abnormalities contribute moderately to heart disease probability. While not the most significant factor, it remains relevant in clinical contexts. A one-unit change in ECG findings leads to a 3.75% point increase in the likelihood of disease, assuming confounders are constant.

##### 8. Max Heart Rate Achieved (thalach)

- ATE: -0.0023
- Confounders: oldpeak, trestbps, chol, age

Interpretation: Maximum heart rate achieved is negatively correlated with heart disease risk, suggesting that better cardiovascular fitness may be protective. Statistically, a one-unit increase in maximum heart rate results in a 0.23% point decrease in heart disease probability, given fixed confounding variables.

##### 9. Exercise-Induced Angina (exang)

- ATE: 0.3598
- Confounders: oldpeak, thalach, trestbps, chol, age

Interpretation: This is one of the most influential features. Patients experiencing angina during exercise are significantly more likely to develop heart disease. The ATE suggests that a one-unit increase in exercise induced angina leads to a 35.98% point increase in heart disease probability, when other confounders are controlled.

##### 10. ST Depression Induced by Exercise (oldpeak)

- ATE: 0.0925
- Confounders: thalach, slope, trestbps, chol, age

Interpretation: ST depression during stress testing moderately increases the risk of heart disease. On average, each unit increase in oldpeak corresponds to a 9.25% point rise in heart disease probability, assuming other variables are fixed.

##### 11. ST Slope (slope)

- ATE: 0.3260
- Confounders: oldpeak, thalach, trestbps, chol, age

Interpretation: The slope of the ST segment is a strong causal indicator of heart disease, particularly in exercise ECG tests. A one-unit increase in the slope of the ST segment leads to a 32.60% point increase in disease probability, controlling for related confounders.

Overall, exercise-induced angina (exang), ST slope, fasting blood sugar, and chest pain type demonstrate the strongest causal effects. Variables traditionally considered important, like cholesterol or resting BP may not have strong causal impact in this dataset, potentially due to mediation or correlation with stronger signals. This causal analysis not only supports known clinical relationships but also justifies the feature selection used in the voting classifier.

## 5 Conclusion

This research presents a comprehensive and ethically aligned AI ecosystem for cardiovascular disease diagnosis to address critical gaps in early detection, clinical applicability, and Responsible AI practices. By integrating advanced tabular neural networks, ensemble models, Bayesian Neural Network, and causal model, robust diagnostic performance is achieved with 0.90 accuracy while ensuring interpretability, uncertainty quantification, and bias mitigation. The work stands out by developing non-clinical models for accessible, early risk assessment, enabling proactive interventions before symptoms manifest. The non-clinical model is designed to be integrated into patient-facing mobile applications or web portals, allowing individuals to assess their risk using self-reported data before seeking specialized medical care. This serves as a filter to prioritize high-risk individuals in resource-constrained healthcare systems. The aim was to enhance transparency, fairness, and trustworthiness in clinical decision-making, by incorporating SHAP, DiCE counterfactuals, FairLearn, and Bayesian uncertainty. Demographic and regional biases are addressed through unsupervised clustering to ensure equitable performance across diverse populations. A novel ML-system (CGEVC) guided by causal inference is also proposed for diagnosing purposes, obtaining an accuracy and precision of 0.89, while reducing dataset dimensionality. Compared to prior works that focus narrowly on accuracy, this approach aligns with FDA Good ML Practices and EU AI ethics, offering a holistic solution for CVD diagnosis. Future directions include expanding datasets for rare CVD subtypes, integrating multimodal data (e.g., ECG, imaging), validating data augmentation procedures in clinical sectors, and refining causal inference for personalized treatment insights. Future research should also focus on reducing disparities in equalized odds while preserving overall model performance. By combining technological innovation with ethical rigor, this research advances AI-driven healthcare toward safer, more equitable, and actionable outcomes.

## Declarations

### Ethical Approval

Not applicable.

#### Funding

The study did not receive any funding.

### Data availability statement

Data used in this research, materials, and reproducibility guidelines are provided in the GitHub repository: https://github.com/SakibHasanSimanto/CVD-AI-Research. Original source of data is UCI Machine Learning Repository: https://archive.ics.uci.edu/dataset/45/heart+disease

